# Development and optimization of self-collected, field stable, saliva-based immunoassays for scalable epidemiological surveillance of pathogen-specific immunity

**DOI:** 10.64898/2026.03.05.26347729

**Authors:** Lauren E Bahr, Joseph Lu, Darunee Buddhari, Taweewun Hunsawong, Erica Rapheal, Peter Greco, Lisa Ware, Michelle Klick, Aaron Farmer, Frank Middleton, Stephen J Thomas, Kathryn Anderson, Adam T Waickman

## Abstract

Serological surveillance is fundamental to infectious disease research and informed public-health decision making. Immunoassays used in the study of pathogen-specific immunity have historically relied on the collection of venous blood. While critical for many public-health applications, this sample collection method is invasive and resource intensive. The costs and logistical barriers associated with venous blood collection are exacerbated in resource-limited regions, and the shift to less invasive sampling methods would increase sample availability for pathogen surveillance and study of pathogen-specific immunity. To this end, we have developed and optimized a self-collected, saliva-based immunoassay capable of quantifying pathogen-specific antibody binding in saliva samples. Using samples collected from geographically and epidemiologically diverse regions of the world, we compared antigen-specific IgG levels in paired plasma and saliva samples. We observed that levels of IgG against multiple pathogens of public health concern – including SARS-CoV-2 and dengue virus (DENV) – were highly correlated in plasma and swab-collected saliva. In addition, the decay of maternally derived antibodies in saliva samples collected from infants was readily observed using this immunoassay, demonstrating the assay’s sensitivity and potential for use in measuring antibody kinetics. We posit that this assay represents a climate stable, non-invasive tool that can aid in the surveillance and study of pathogen-specific immunity across a broad range of public-health indications.

## INTRODUCTION

Serological surveillance is a cornerstone of infectious disease epidemiology, informing critical public health decisions and offering insight into both historic and ongoing patterns of pathogen transmission. Traditionally, this analysis has relied on serology-requiring venous blood collection and a subsequent plasma or serum isolation for use in assays such as ELISA, *in vitro* neutralization assays, and multiplex antigen arrays ^1^. While serum and plasma are robust and reliable sources of pathogen-specific antibodies, venous blood collection from infants, young children, elderly individuals, and individuals experiencing acute illness can be logistically challenging. There is also a limit to how often blood can be collected, even from healthy adult volunteers, as trained phlebotomists are required and study participants may be required to consent to repeated draws. Finally, blood samples must be stored and processed correctly in a temperature and time sensitive manner. This is also a costly endeavor, requiring trained staff for sample collection, transportation, and processing within a timeline that maximizes sample stability.

High-frequency and longitudinal sampling is often essential for accurate serological interpretation and/or detection of a recent pathogen infection^2–4^. These challenges are especially acute for antigenically related pathogens such as orthoflaviviruses, where extensive immunologic cross-reactivity between dengue virus (DENV) serotypes, Zika virus (ZIKV), Japanese encephalitis virus (JEV), and yellow fever virus (YFV) can confound interpretation of exposure history and immune status. Quantifying the development and evolution of these complex and cross-reactive immune profiles and evaluating how they relate to risk of infection and/or severe disease remains challenging, especially with the logistical limitation imposed by blood-based sample collection.

Recent strides have been made in the development of saliva-based assays, with the global SARS-CoV-2 pandemic expediting the development of saliva-based PCR and rapid antigen tests^5,6^. Saliva-based assays for detecting HIV^7,8^, certain cancer markers^9,10^, dental conditions^11^, and bacteria that can lead to stomach ulcers^12^ have been developed in recent years. Saliva sample collection is minimally invasive, requires minimal training of study team personnel, can be performed at home by the volunteer, and can be stored in a stabilizing buffer solution at ambient temperature, allowing for more time between sample collection and sample processing without impacting sample quality. Both pathogen and host RNA and DNA can be extracted from saliva samples, and IgG and IgA antibody response can be readily measured. Salivary IgG closely matches the repertoire of serum IgG, albeit at a lower concentration^13^. Notably, a swab-based collection method can be used in which saliva is preferentially collected from the crevicular fluid, where salivary IgG best reflects serum IgG^14^.

In this study, we systematically evaluate a self-administered, cold-chain-free, swab-based saliva collection strategy targeting crevicular fluid for quantifying pathogen-specific IgG responses. To this end we collected paired plasma and saliva samples and compared the total IgG and antigen-specific IgG levels across geographically and epidemiologically distinct cohorts. This analysis revealed a high degree of correlation between pathogen-specific IgG titers in plasma and saliva, which was stable over time and reflected anticipated changes elicited by infection and/or vaccination events. Additionally, we observed maternal antibody decay in saliva samples from mother/infant dyad pairs. These results indicate that saliva-based assays are capable of measuring waning immunity where traditional blood-based assays have been less sensitive. These results suggest that saliva-based immunoassays are a durable, easily deployable, and sensitive platform for seroepidemiological research that can be readily used in resource-limited research environments.

## RESULTS

### Assessment of saliva as a source of pathogen-specific IgG

The first objective of this study was to evaluate the quantity, specificity, and stability of salivary IgG collected using a self-administered swab-based approach. To this end, volunteers were enrolled in an initial validation study in Syracuse, NY. Upon enrollment, a plasma and saliva sample was collected from each subject in a clinical setting, and participants were instructed on how to perform a self-administered saliva collection and provided additional saliva collection kits for home-collection. Saliva swabs were subsequently collected daily for four consecutive days and stored at room temperature. After four days, subjects mailed the set of four tubes to a central lab for processing and analysis.

Consistent with prior reports, total IgG levels in enrollment saliva samples were substantially lower – yet consistent - than in paired plasma samples (**Supplemental Figure 1**). Having confirmed that human IgG could be reliably measured using this swab-based collection method, antigen-specific IgG concentrations were compared in paired plasma and saliva samples. IgG binding to spike, receptor-binding domain (RBD), and nucleocapsid proteins from SARS-CoV-2 were measured using a multiplexed immunoassay. Significant positive correlations were observed between antigen-specific IgG in paired plasma and saliva samples for all three antigens (Pearson=0.867, Spearman=0.81)(**Figure 1A**). To account for the lower total IgG concentration in saliva, correlations were also performed using saliva data normalized to total IgG concentration. These correlations remained significantly positive, (Pearson=0.869, Spearman=0.824), indicating that despite lower overall IgG levels, antigen-specific antibodies were robustly represented in salivary samples (**Figure 1B**).

**Figure 1.**
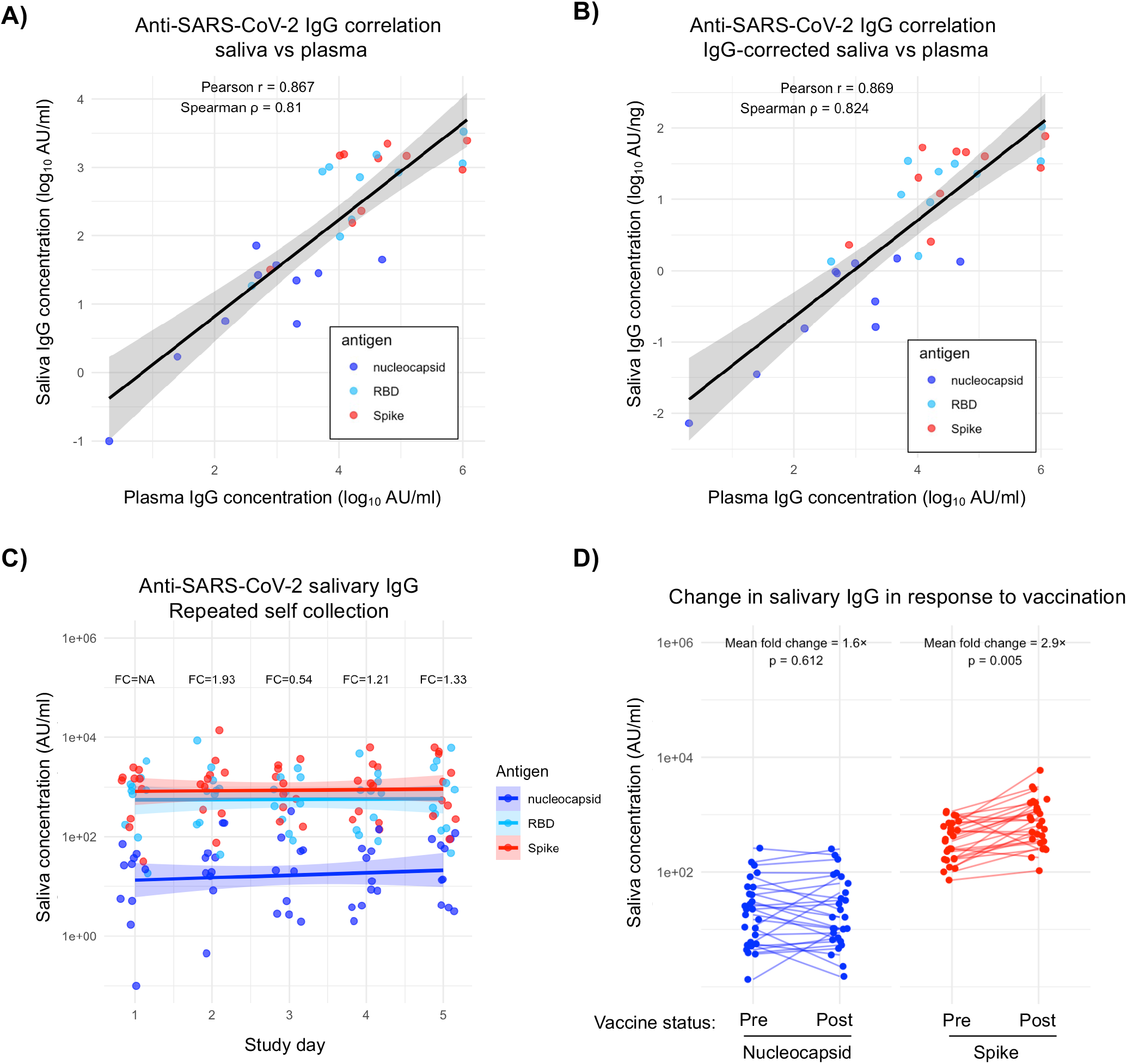
Saliva as a source of pathogen-specific IgG. **A)** Pearson’s and Spearman’s correlation between SARS-CoV-2 antigen specific plasma and saliva IgG values uncorrected for total salivary IgG concentration. Spline with shaded region indicating 95% confidence interval. **B)** Pearson’s and Spearman’s correlation between SARS-CoV-2 antigen specific plasma and saliva IgG values corrected for total salivary IgG concentration. Spline with shaded region indicating 95% confidence interval. **C)** Measure of changes in daily antigen-specific salivary IgG concentrations. Splines of the datapoints shown for each antigen, with the shaded regions representing a 95% confidence interval. FC indicates fold changes from previous day’s average. **D)** Measure of antigen-specific IgG levels in pre- and post-SARS-CoV-2 vaccination saliva samples. Nucleocapsid and spike reactive IgG measured and mean fold change for each antigen calculated.

A key objective for saliva-based seroepidemiology is enabling high-frequency longitudinal sampling—for example, daily collection following infection to capture acute antibody kinetics. To evaluate the consistency of self-collected samples over time, we quantified the abundance of SARS-CoV-2 reactive salivary IgG in the daily self-collected saliva swabs taken over the course of 5 days. While minor day-to-day variations were observed, all fluctuations remained under 2-fold throughout the study period, indicating relatively stable and consistent results with repeated sampling (**Figure 1C, Supplemental Figure 2**).

Having demonstrated the day-to-day stability of pathogen-specific IgG in the absence of infection or vaccination, we next sought to determine if salivary IgG levels change measurably in response to antigen exposure. To this end, we utilized samples from a respiratory virus surveillance study at SUNY Upstate Medical University, in which employees provided saliva samples at enrollment and before and after annual SARS-CoV-2 vaccination. We analyzed the paired pre- and post-vaccination saliva samples for levels of anti-nucleocapsid and anti-spike protein IgG (**Figure 1D**). Consistent with the day-to-day variation observed with saliva self-collection in the absence of vaccination or infection, we found that the mean fold change in anti-nucleocapsid antibody levels remained under 2-fold pre/post-vaccination (paired t-test p=0.612), as expected for a vaccine that doesn’t contain nucleocapsid antigen. In contrast, there was a 2.9- fold increase in the abundance of anti-spike salivary IgG after vaccination (paired t-test p=0.005), indicating that the vaccine induced an anti-spike protein immune response that was measurable in human salivary samples. Overall, these data indicate that salivary IgG is antigen-specific in a manner comparable to IgG in plasma, stable upon repeated self-collection, and changes in response to antigen exposure.

### Assessment of saliva as a source of pathogen-specific IgG in a multigenerational household cohort study

In light of the results of our preliminary analysis, we next sought to evaluate the suitability of saliva-based immunologic testing in an epidemiological survey. We utilized specimens collected from a longitudinal, multigenerational family cohort study based in Kamphaeng Phet, Thailand. In this study multigenerational families are enrolled beginning with a pregnant mother. Blood and saliva samples are taken from the family upon enrollment, and cord and maternal blood are collected at birth if available alongside saliva samples from the mother and the infant. Additional blood and saliva samples are collected every 6 months, and a series of saliva and blood samples are taken from the entire household if a family member experiences a febrile illness. With these samples we were able to directly compare the performance of saliva-based assays to well-established plasma-based assays in the field and across a wide range of ages.

To bridge our initial validation study to this family cohort study, we first quantified the abundance of SARS-CoV-2 specific IgG in both plasma and saliva samples from individuals aged 0 to 80, an age range representative of the general cohort population. In analysis of saliva samples not corrected for total IgG concentration, the levels of antigen-specific salivary IgG strongly correlated (Pearson=0.745, Spearman=0.742) with that of plasma IgG (**Figure 2A**). This strong correlation was maintained (Pearson=0.748, Spearman=0.762) when the saliva data were corrected for total IgG concentration (**Figure 2B**). To better understand what effect, if any, volunteer age had on the quality of salivary IgG collected in this analysis, we performed a sliding window correlation analysis (**Figure 2C**). The correlation between saliva and plasma SARS-CoV-2 specific IgG titers did not vary significantly with age, and this relationship was not significantly impacted by correcting for the total IgG content in saliva sample (Fisher’s Z test p=0.872). Trends in absolute levels of salivary IgG against nucleocapsid, RBD, and spike antigens by age were observed, with anti-spike and RBD IgG antibodies increasing from birth to 25 years of age (**Figure 2D**). In contrast, stable levels of anti-nucleocapsid IgG were observed across age groups. These observations are consistent with vaccination eliciting antibodies against spike and RBD antigens. This same trend holds true when the antigen-specific IgG data is corrected for total IgG concentration (**Supplemental Figure 3**). These data indicate that saliva samples can reliably be used to test for antigen-specific IgG across a broad range of subject ages.

**Figure 2.**
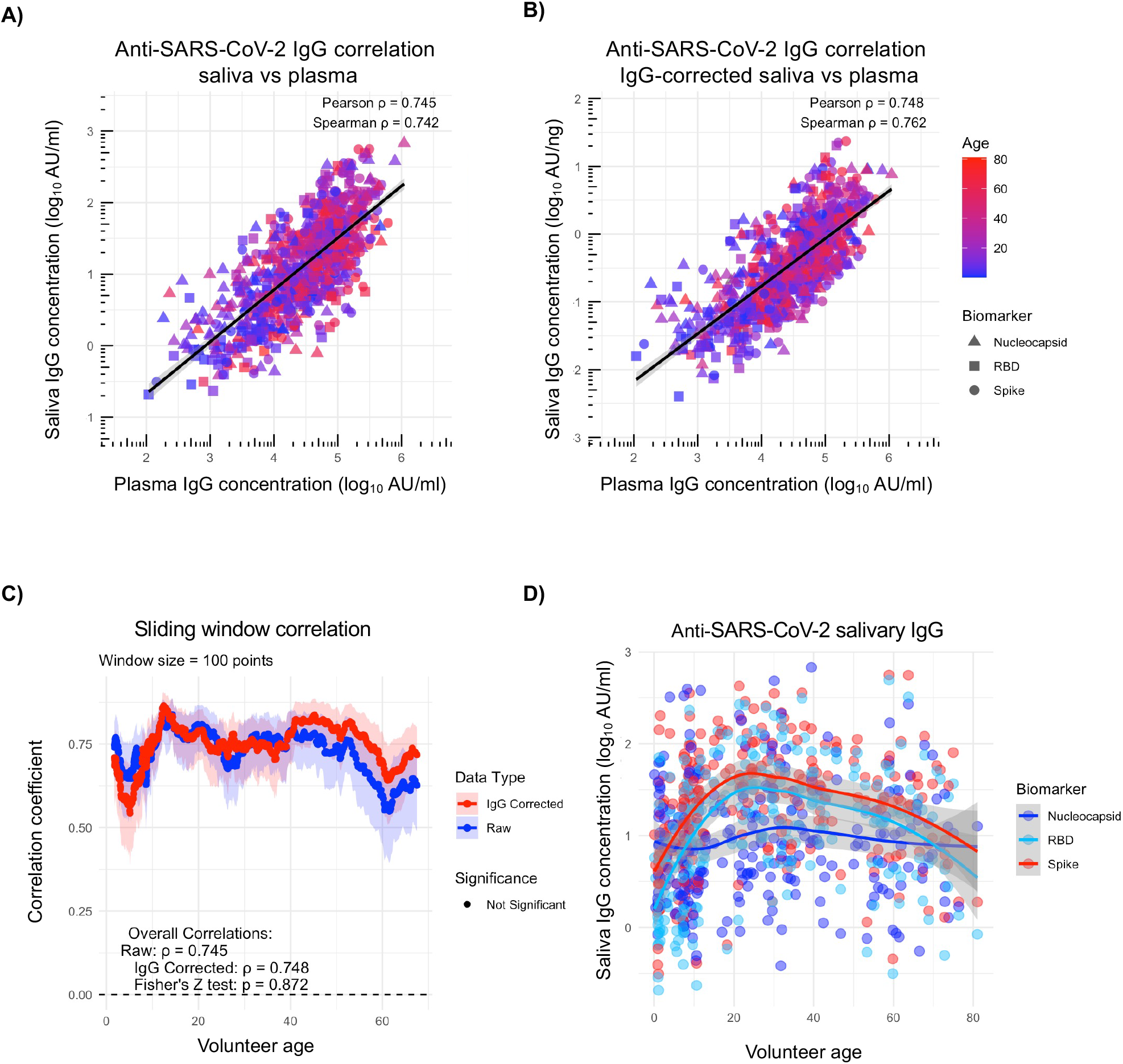
Saliva as a source of pathogen-specific IgG in a multigeneration household cohort study. **A)** Correlation between plasma and saliva anti-nucleocapsid, RBD, and spike IgG values uncorrected for total salivary IgG concentration. Spline with shaded region indicating 95% confidence interval. **B)** Correlation between plasma and saliva anti-nucleocapsid, RBD, and spike IgG values corrected for total salivary IgG concentration. Spline with shaded region indicating 95% confidence interval. **C)** Sliding window correlation comparing age and correlation coefficient for both raw (uncorrected) and IgG corrected salivary data. **D)** Plot of saliva anti-nucleocapsid, RBD, and spike IgG values uncorrected for total salivary IgG concentration by subject age. Saliva IgG concentrations plotted on a log scale. Point and spline colored by antigen. Spline plotted for each antigen, shaded region represents 95% confidence interval.

### Saliva as a source of DENV-specific IgG

Having confirmed the ability of saliva collection to capture changes in antibody levels consistent with a respiratory virus antigen exposure, we next sought to expand these observations to DENV-specific immunity. Accordingly, we developed a pan-DENV IgG assay, utilizing an equimolar pool of DENV-1, -2, -3, and -4 NS1 antigen. Using this assay and a subset of samples from the Kamphaeng Phet family cohort study, we observed a significant correlation between anti-DENV salivary and plasma IgG levels in data both uncorrected for total salivary IgG concentration (Pearson=0.917, Spearman=0.908) (**Figure 3A**) and corrected for total salivary IgG concentration (Pearson=0.93, Spearman=0.921) (**Figure 3B**). A sliding window correlation on this data demonstrated that correcting the values for total IgG did not increase the statistical significance of this relationship across a broad age range (uncorrected for total IgG Pearson=0.917, corrected for total IgG Pearson=0.93) (**Figure 3C**).

**Figure 3.**
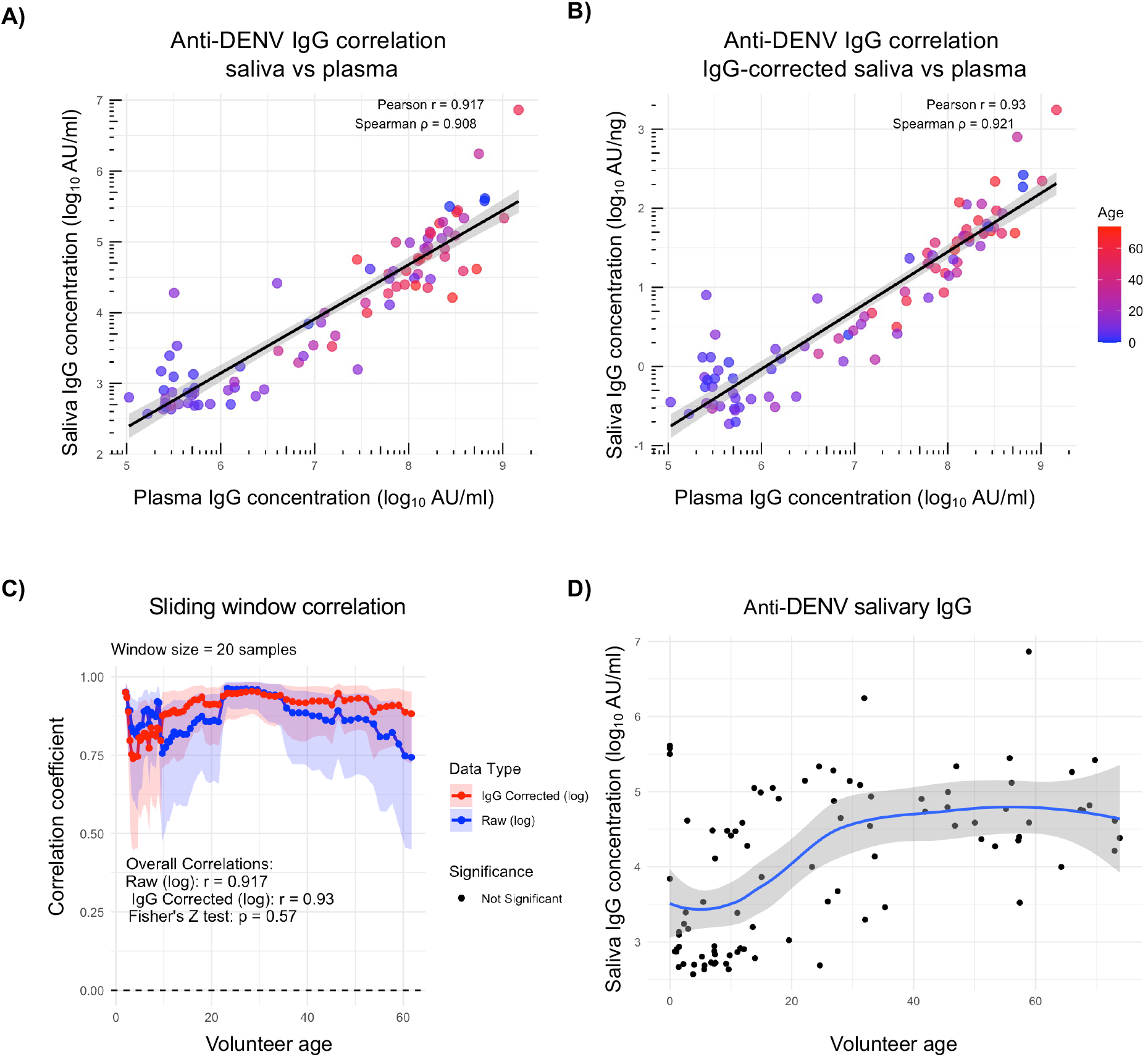
Saliva as a source of DENV-specific IgG. **A)** Correlation between plasma and saliva anti-DENV 1-4 NS1 IgG values uncorrected for total salivary IgG concentration. Plasma and saliva values plotted on log10 scale. **B)** Correlation between plasma and saliva anti-DENV 1-4 NS1 IgG values corrected for total salivary IgG concentration. **C)** Sliding window correlation comparing age and correlation coefficient for both raw (uncorrected) and IgG corrected salivary data. **D)** Correlation between saliva anti-DENV 1-4 NS1 IgG values uncorrected for total salivary IgG concentration and subject age. Spline shown in blue and shaded region represents 95% confidence interval.

Consistent with the known epidemiology of DENV in Kamphaeng Phet, Thailand, we observed an increase in DENV-specific IgG levels with age in individuals enrolled in this cohort (**Figure 3D, Supplemental Figure 4**)^15^. These data support the use of saliva samples to quantify anti-DENV antibodies and capture changes in these antibody levels associated with age in addition to exposure.

### Saliva IgG collection captures maternal antibody decay in infants

Having validated that antibody responses can be measured in samples from subjects of all ages, we wanted to investigate the capability for saliva samples to capture maternal antibody decay. The accurate quantification of maternally derived antibodies is of particular interest for DENV, as waning maternal antibody titers have been associated with an increased risk of severe dengue in infants^16,17^. Using an immunoassay to measure IgG levels against SARS-CoV-2 antigens and DENV 1-4 NS1 in saliva samples from mother-infant dyads, we observed that the levels of antigen-specific IgG in saliva of mother/infant dyads were highly correlated at birth, consistent with the maternal origin of the infant antibodies in cord blood (**Figure 4A**). To gain further insight into the kinetics of maternal antibodies in infant saliva samples we quantified antigen-specific IgG levels in the infant saliva samples in the first 4-5 months after birth and observed that these antibody levels decayed substantially over time, in a pattern and rate consistent with loss of maternal antibodies after birth (**Figure 4B**). In contrast, the level of these same antibodies remained constant in the mothers of these infants (**Supplemental Figure 5**). This further indicates that we are able to quantify maternal antibody decay using saliva samples from infants that were collected at home.

**Figure 4.**
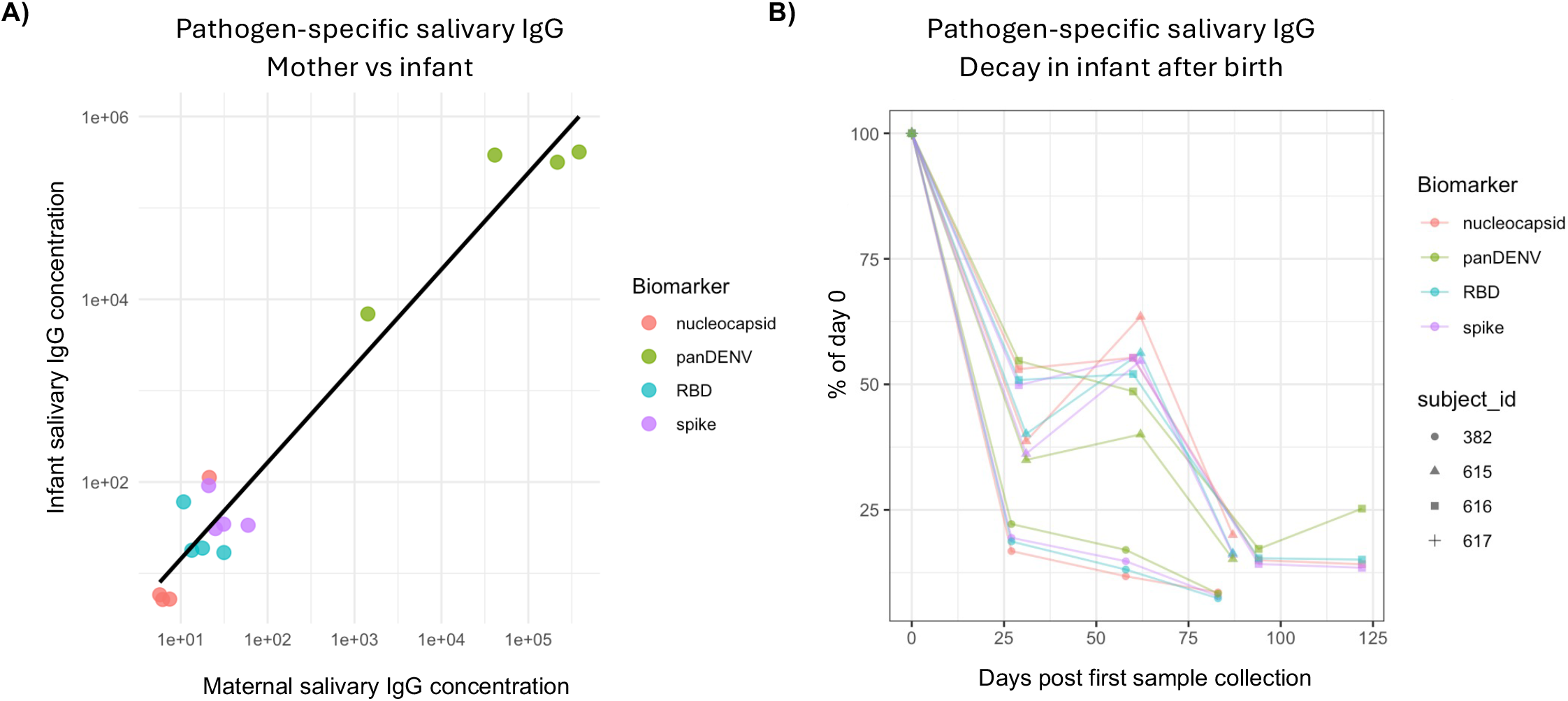
Salivary IgG collection captures maternal antibody decay in infants. **A)** Plot of antigen-specific salivary IgG levels in samples from paired mother and infant saliva samples. Samples uncorrected for total IgG concentration. Linear regression represented by black line. **B)** Decay of maternal antibodies in infant salivary samples following birth. Infant salivary IgG data from first sample normalized to 100 and decay plotted as a function of percent of enrollment sample.

## DISCUSSION

Saliva-based immunoassays offer a promising alternative to traditional blood-based serological assays, addressing several persistent barriers to large-scale and longitudinal seroepidemiological research. The data presented herein demonstrate that a self-administered (or adult-assisted collection in the case of small children), swab-based saliva collection strategy targeting crevicular fluid can reliably quantify pathogen-specific IgG responses across diverse populations and settings.

The use of saliva as a diagnostic tool is appealing on many fronts. It is minimally invasive, in our case only requiring the subject to place a sponge-like swab in their cheek near the gums for two minutes. It is more cost, time, and labor effective than blood samples, which require trained staff with sterile equipment and often rely on either clinical staff or enrolled subjects travelling for sample collection. Blood sample integrity can be difficult to maintain in certain climates, and factors such as temperature and time until sample processing can impact sample quality. In regions endemic for neglected tropical diseases the resources and infrastructure needed for blood sampling and processing are not always available. Saliva as an alternative platform for sampling does not require staff or clinical visits since self-collected or parent collected samples can be stabilized in a buffer to be stored within a wide ambient temperature range, and repeated samples can be obtained comfortably, approximately every 2 hours, with the only caveats being consumption of recent food or drink, use of gum, lozenges or tobacco products, and performance of oral hygiene.

The ability to perform frequent, non-invasive sampling opens up many possibilities for study design moving forward. The agnostic nature of this platform allows for customization of target antigen. In the case of orthoflaviviruses like DENV, frequent sampling may help capture subclinical infections which make up the bulk of DENV infections globally^18^. The kinetics and effect of subclinical DENV infections are still not fully understood, but these infections contribute to DENV circulation within a population and are important to consider when designing a vaccine^19,20^. Mathematical modeling of anti-DENV antibody decay rates suggests sampling at 3-month intervals increases the likelihood of capturing subclinical infections^21^. Obtaining blood samples at this frequency is difficult but shifting towards less invasive at home methods of sample collection, such as saliva, may fill this gap in available data. The quantification of DENV-specific immunity is complicated by the co-circulation of multiple DENV serotypes and antigenically related viruses (such as ZIKV, JEV, and YFV) in many regions of the world. Accordingly, inferring recent DENV infection by retrospective serology is challenging and best achieved with frequent sample collection, such as that enabled by saliva analysis. This saliva platform can be applied to other pathogens, perhaps even in a rapid test form. Currently a plasma based DENV NS1 antigen rapid test is on the market, and developments in saliva platform such as the one described in this submission may lend itself to diagnostic innovations.

It must be noted that this study is far from the first to describe a method for quantifying the abundance of DENV-specific IgG in saliva in an effort to bypass the need for venous blood collection^22–24^. These prior studies have utilized both expectoration and swab-based collection methodologies^23,25^. However, these prior studies have demonstrated only modest correlation with blood-based methodologies, especially when attempting to identify historic orthoflavivirus infections^26^. We posit that the methodology outlined in this report significantly advances upon these prior studies due to the combined use of crevicular fluid focused collection swabs with a protein-stabilization solution - which significantly improves the recovery and stability of IgG after collection – as well as a high-sensitivity endpoint assay that is able to compensate for the lower concentration of IgG found in saliva versus plasma or serum.

A limitation of this study is the recent implementation of saliva sample collection in the Kamphaeng Phet family cohort study, resulting in a limited number of saliva samples being available for analysis. The same limitation applies to the mother/infant dyads analysis, as more dyads are enrolled these monthly samples will continue. Notably, an important consideration of sampling in mother/infant dyads is the time since nursing, since maternal antibodies can be introduced directly to the oral cavity from breast milk. Therefore, caution must be taken if examining isotypes other than IgG. In this study, however, staff assisted with the collection of saliva from mother/infant dyads to control for these variables. Additionally, from a technical perspective, the stabilization media used to collect and store the saliva samples is incompatible with living cells. Accordingly, the salivary content collected as described in this study is incompatible with cell-based neutralization assays and/or any assay necessitating the use of living cells.

In conclusion, we posit that saliva-based immunoassays represent a low-cost and non-invasive alternative to traditional blood-based assays for the quantification and longitudinal assessment of anti-pathogen immunity, and that the methodology outlined here represents a significant advancement in our ability to successfully implement this promising technology. In the future we hope to continue using this saliva platform to investigate orthoflavivirus immunity, with a focus on subclinical DENV infections. We also plan to expand the immunoassay to include co-circulating and related orthoflavivirus such as Zika virus (ZIKV) and Japanese encephalitis virus (JEV) to investigate the impact of sequential infections and immune history on subsequent infections and vaccinations.

## METHODS

### Saliva collection and processing

Saliva for this study was collected using an OraSure Oral Fluid Collection Device (OraSure, 503-0507). Volunteers were asked not to eat, drink, or breastfeed infants (for infant saliva sample) prior to collection. Samples were collected using a collection pad that subjects place in the crevicular space between the gum and lower cheek for 2-5 minutes. The collection pad is then broken off into the specimen vial of stabilizing buffer for storage until sample processing. During processing breakaway tabs on the bottom of specimen tubes are removed before placing the specimen tube into a conical tube and centrifuging to collect sample. Samples were stored at -80° C until analysis.

### Plasma collection and processing

Venous blood was collected in EDTA tubes and processed within 16 hours of collection. Whole blood was centrifuged to collect plasma, and samples were stored at -80° C until analysis.

### COVID Employee Surveillance Study

Healthcare workers at SUNY Upstate Medical University were enrolled in a prospective cohort study to monitor SARS-CoV-2 infection and vaccine immunogenicity during the 2024-2025 respiratory virus season. Saliva samples were collected at enrollment (pre-season) and at the end of the season (post-season) for antibody testing using the protocol outlined above. Participants who experienced COVID-like illness contacted the study team and self-collected daily saliva swabs for five consecutive days beginning at symptom onset for PCR testing, viral culture, and sequencing. Participants who received a COVID-19 vaccine during the study period provided pre- and post-vaccination saliva samples for antibody analysis. This study and the associated analysis were approved by the Institutional Review Board for the Protection of Human Subjects, State University of New York Upstate Medical University, with written and informed consent was obtained from all study volunteers.

### Kamphaeng Phet family cohort study

As previously described^27^ this longitudinal study consists of multigenerational families within the same household including a pregnant mother as well as another child and a family member from the generation above the mother. Blood and saliva samples are collected upon enrollment, at newborn delivery, and semi-annually. If an individual in the family has a confirmed DENV infection the entire family gives three illness investigation blood and saliva samples at acute and convalescent timepoints. Saliva samples were collected from mothers and their infants monthly starting at the delivery of the infant. Saliva and plasma samples from enrollment and mother-infant dyad saliva samples were used in this study. This study was approved by the Institute for the Development of Human Research Protections, Thailand Ministry of Public Health; University Research Ethics Committee, University of Cambridge; Institutional Review Board for the Protection of Human Subjects, State University of New York Upstate Medical University; and Walter Reed Army Institute of Research Institutional Review Board. Written informed consent or assent was obtained from all study volunteers.

### Total human IgG capture ELISA

Human IgG Total Uncoated ELISA Kit (Invitrogen, REF 88-50550-88) was used to measure total IgG levels in saliva samples. Saliva samples diluted 1:50 and further diluted 1:200 if out of assay range.

### SARS-CoV-2 immunoassay using Meso Scale Discovery (MSD) platform

SARS-CoV-2 immunoassay kits from MSD (Catalog #K15383U-2) were used to measure the levels of IgG that bound to spike, nucleocapsid, and RBD antigens that had been pre-coated on a 96-well plate. Plates were blocked at room temperature shaking at 700 rpm for 30 minutes with MSD Blocking Buffer A. Plates were washed with MSD Wash Buffer (1X) and then standard dilutions, controls, and plasma or saliva samples previously diluted in MSD Diluent 100 were added to the plate and incubated at room temperature shaking at 700 rpm for 2 hours. Plasma samples diluted 1:10,000, saliva samples diluted 1:50. Plates were washed then coated with anti-IgG secondary antibody and incubated at room temperature shaking at 700 rpm for 1 hour. Plates were washed one final time and then coated with MSD Gold Read Buffer B and results were read on an MSD instrument. Raw data reported as electrochemiluminescent values, data was exported and analyzed for concentration values using MSD software. Further analysis done using R and RStudio.

### DENV 1-4 NS1 immunoassay using Meso Scale Discovery (MSD) platform

Custom immunoassays were created using a U-Plex Development Pack (7-assay) from Meso Scale Discovery (MSD, Catalog #K15232N-4). This assay allows for up to 7 unique biotinylated antigens to coat each well of a 96-well plate. In this assay recombinant DENV-1 NS1, DENV-2 NS1, DENV-3 NS1, and DENV-4 NS1 (Native Antigen) were biotinylated and then mixed at a 1:1:1:1 ratio before being conjugated to an MSD linker protein which allows the antigens to coat the plate. 96-well MSD plates were coated with antigen conjugated with linker at room temperature shaking at 700 rpm for 1 hour. Plates were washed with MSD Wash Buffer (1X) then blocked at room temperature shaking at 700 rpm for 30 minutes with MSD Blocking Buffer A. Plates were washed, then plasma or saliva samples that had been diluted in MSD Diluent 100 were added to the plate and incubated at room temperature shaking at 700 rpm for 1 hour. Plasma samples diluted 1:1000, saliva samples diluted 1:5. Plates were washed then coated with anti-IgG secondary antibody and incubated at room temperature shaking at 700 rpm for 1 hour. Plates were washed one final time and then coated with MSD Gold Read Buffer B and results were read on an MSD instrument. Raw data reported as electrochemiluminescent values, with further analysis performed using R and RStudio.

### Statistical analysis

Statistical analysis and data visualization was performed using R Statistical Software (R version 4.5.2), RStudio (version 2025.09.2+418).

## Acknowledgements

We gratefully acknowledge the members of the Global Health Institute (GHI) of SUNY Upstate Medical University and the members of WRAIR-AFRIMS in Bangkok and Kamphaeng Phet for their support of these studies. Material has been reviewed by the Walter Reed Army Institute of Research. There is no objection to its presentation and/or publication. The opinions or assertions contained herein are the private views of the author, and are not to be construed as official, or as reflecting true views of the Department of the Army or the Department of Defense. The investigators have adhered to the policies for protection of human subjects as prescribed in AR 70–25.

## Data Availability Statement

All data required for the reproduction of the analysis presented in this study are present in the manuscript or in the corresponding supplemental data files.

## REFERENCES

1. Hamins-Puértolas, M. et al. Linking multiple serological assays to infer dengue virus infections from paired samples using mixture models. PLoS Comput. Biol. 21, e1013708 (2025).

2. Conrey, S. C. et al. Optimizing the Sensitivity of Detection of Respiratory Syncytial Virus Infections in Longitudinal Studies Using the Combination of Weekly Sample Testing and Biannual Serology HHS Public Access.

3. Henderson, A. D. et al. Zika seroprevalence declines and neutralizing antibodies wane in adults following outbreaks in french polynesia and fiji. Elife 9, (2020).

4. Haselbeck, A. H. et al. Serology as a Tool to Assess Infectious Disease Landscapes and Guide Public Health Policy. Pathogens vol. 11 Preprint at 10.3390/pathogens11070732 (2022).

5. Miller, E. W. et al. Development and Validation of Two RT-qPCR Diagnostic Assays for Detecting Severe Acute Respiratory Syndrome Coronavirus 2 Genomic Targets across Two Specimen Types. Journal of Molecular Diagnostics 24, 294–308 (2022).

6. Thomas, A. C. et al. Evaluation and deployment of isotype-specific salivary antibody assays for detecting previous SARS-CoV-2 infection in children and adults. Communications Medicine 3, (2023).

7. Greenwald, J. L., Burstein, G. R., Pincus, J. & Branson, B. A Rapid Review of Rapid HIV Antibody Tests. http://www.hret.org/hret/programs/hivtransmrpd.html. (2006).

8. Roehr, B. FDA approves ‘instant’ HIV home test. BMJ (Clinical research ed.) vol. 345 Preprint at 10.1136/bmj.e4636 (2012).

9. Wang, Y. et al. Detection of somatic mutations and HPV in the saliva and plasma of patients with head and neck squamous cell carcinomas. Sci. Transl. Med. 7, (2015).

10. Zhang, L. et al. Salivary Transcriptomic Biomarkers for Detection of Resectable Pancreatic Cancer. Gastroenterology 138, (2010).

11. Zhang, C. Z. et al. Saliva in the diagnosis of diseases. International Journal of Oral Science vol. 8 133–137 Preprint at 10.1038/ijos.2016.38 (2016).

12. Yu, M., Zhang, X. Y. & Yu, Q. Detection of oral helicobacter pylori infection using saliva test cassette. Pak. J. Med. Sci. 31, 1192–1196 (2015).

13. Hettegger, P. et al. High similarity of IgG antibody profiles in blood and saliva opens opportunities for saliva based serology. PLoS One 14, (2019).

14. Tew, J. G., Marshall, D. R., Burmeister, J. A. & Ranney, R. R. Relationship between Gingival Crevicular Fluid and Serum Antibody Titers in Young Adults with Generalized and Localized Periodontitis. INFECTION AND IMMUNITY https://journals.asm.org/journal/iai (1985).

15. Hamins-Puértolas, M. et al. Household immunity and individual risk of infection with dengue virus in a prospective, longitudinal cohort study. Nat. Microbiol. 9, 274–283 (2024).

16. Driscoll, M. O. et al. Maternally derived antibody titer dynamics and risk of hospitalized infant dengue disease. Proc. Natl. Acad. Sci. U. S. A. 120, (2023).

17. Fowler, A. M. et al. Maternally Acquired Zika Antibodies Enhance Dengue Disease Severity in Mice. Cell Host Microbe 24, 743-750.e5 (2018).

18. Bhatt, S. et al. The global distribution and burden of dengue. Nature 496, 504–507 (2013).

19. Duong, V. et al. Asymptomatic humans transmit dengue virus to mosquitoes. Proc. Natl. Acad. Sci. U. S. A. 112, 14688–14693 (2015).

20. Salje, H. et al. Evaluation of the extended efficacy of the Dengvaxia vaccine against symptomatic and subclinical dengue infection. Nat. Med. 27, 1395–1400 (2021).

21. Salje, H. et al. Reconstruction of antibody dynamics and infection histories to evaluate dengue risk. Nature 557, 719–723 (2018).

22. Wasik, D., Mulchandani, A. & Yates, M. V. Salivary Detection of Dengue Virus NS1 Protein with a Label-Free Immunosensor for Early Dengue Diagnosis. Sensors 18, 2641 (2018).

23. Cuzzubbo, A. J. et al. Detection of Specific Antibodies in Saliva during Dengue Infection. JOURNAL OF CLINICAL MICROBIOLOGY vol. 36 (1998).

24. Andries, A. C. et al. Value of Routine Dengue Diagnostic Tests in Urine and Saliva Specimens. PLoS Negl. Trop. Dis. 9, (2015).

25. Anders, K. L. et al. An evaluation of dried blood spots and oral swabs as alternative specimens for the diagnosis of dengue and screening for past dengue virus exposure. American Journal of Tropical Medicine and Hygiene 87, 165–170 (2012).

26. Andries, A. C. et al. Evaluation of the performances of six commercial kits designed for dengue NS1 and anti-dengue IgM, IgG and IgA detection in urine and saliva clinical specimens. BMC Infect. Dis. 16, (2016).

27. Anderson, K. B. et al. An innovative, prospective, hybrid cohort-cluster study design to characterize dengue virus transmission in multigenerational households in Kamphaeng Phet, Thailand. Am. J. Epidemiol. 189, 648–659 (2020).

