## Supplemental figures for "Development and optimization of self-collected, field stable, saliva-based immunoassays for scalable epidemiological surveillance of pathogen-specific immunity"

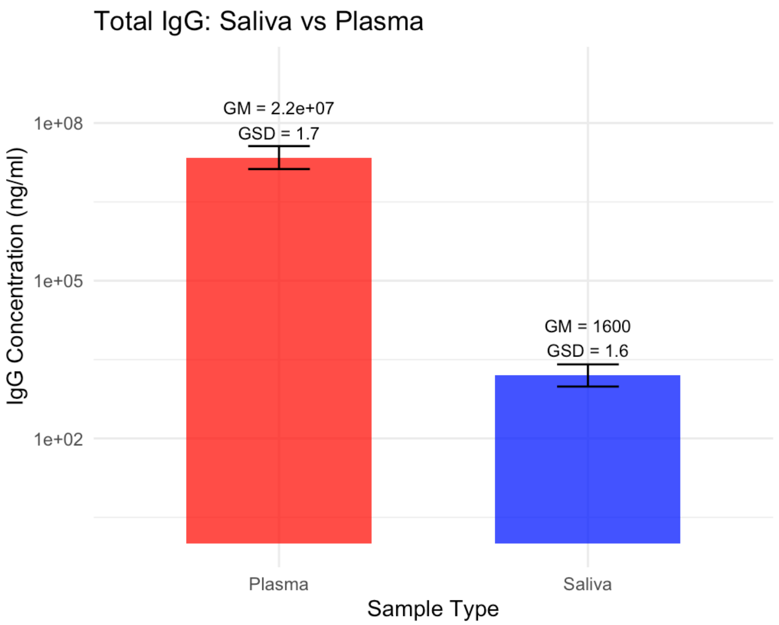


**Supplemental Figure 1.** Concentration of total IgG in serum and saliva as determined by ELISA

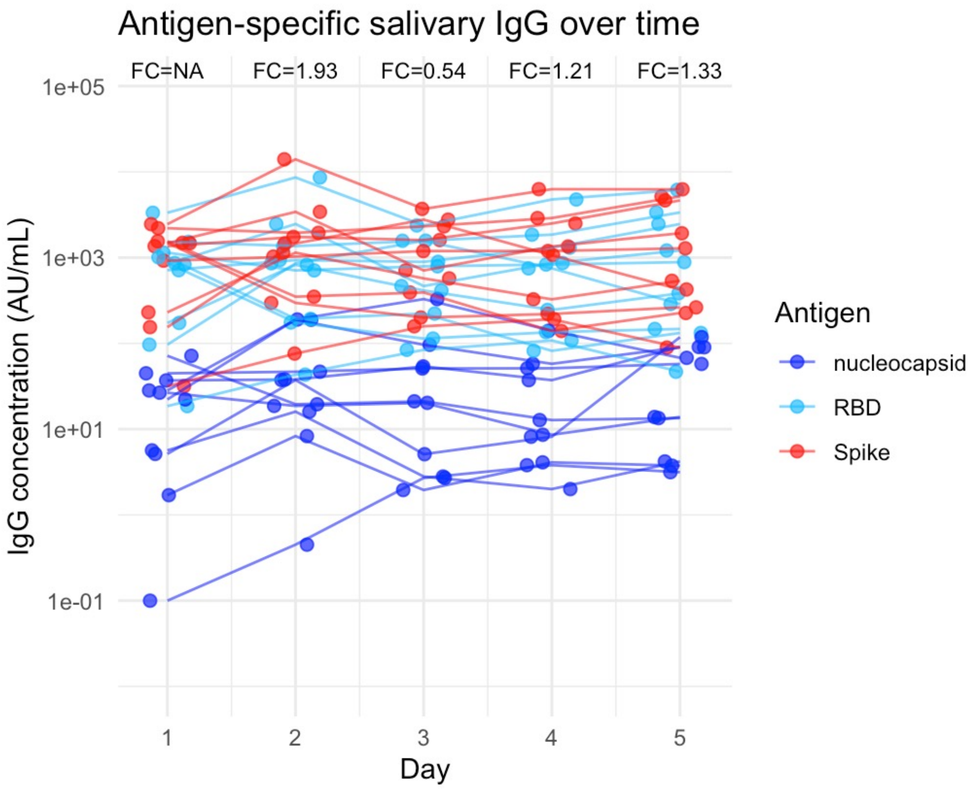


**Supplemental Figure 2.** Concentration of SARS-CoV-2 specific IgG in saliva collected over 5 consecutive days. Lines connect samples connected from the same individual on sequential days

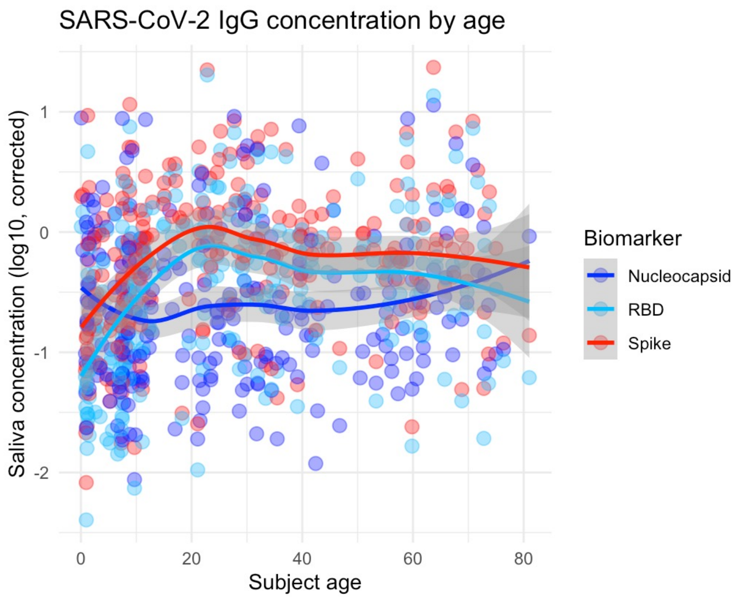


**Supplemental Figure 3.** Concentration of SARS-CoV-2 specific IgG in saliva corrected for total IgG concentration in saliva plotted against volunteer age

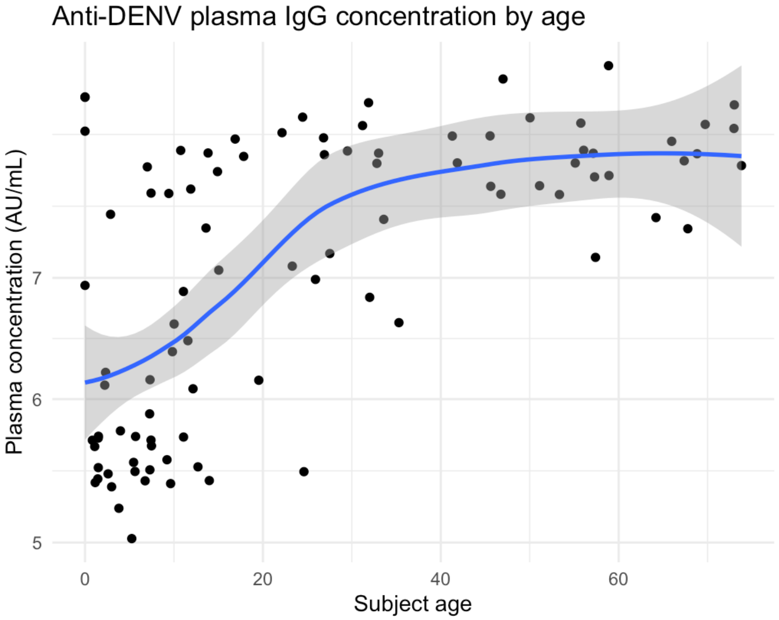


**Supplemental Figure 4.** Concentration of DENV NS1-specific IgG in plasma plotted against volunteer age

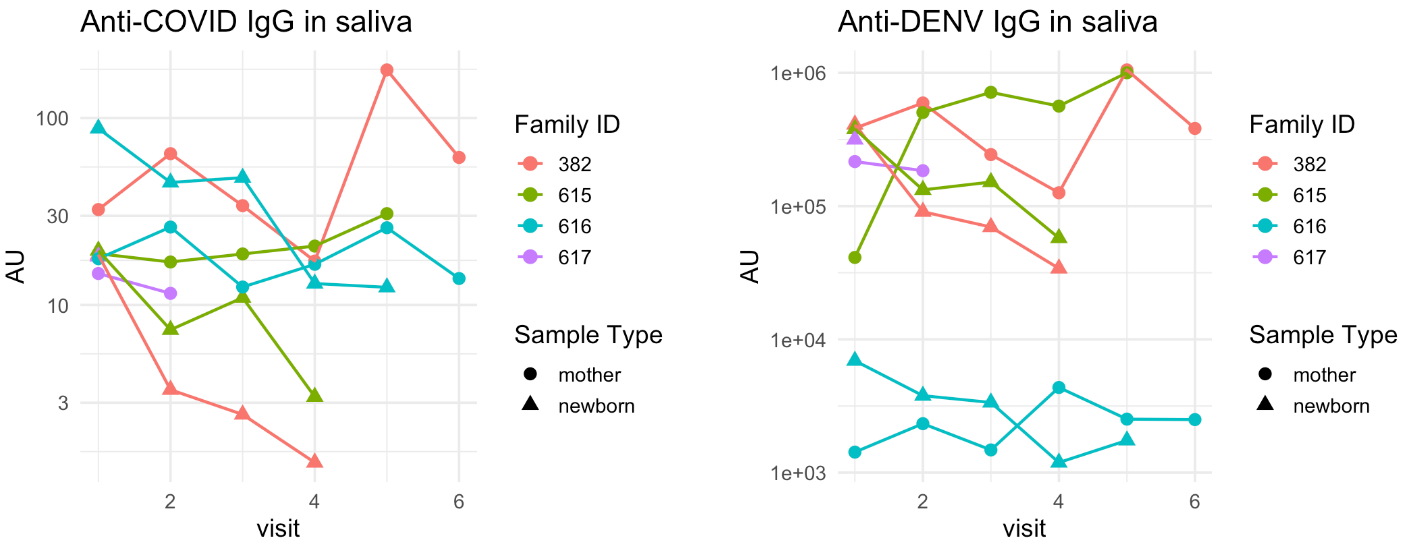


**Supplemental Figure 5.** Concentration of DENV NS1-specific IgG in collected from four mother/infant dyads plotted against the visit from which the same was collected
